# DNA methylation differences at birth after conception through assisted reproductive technologies

**DOI:** 10.1101/2020.03.16.20037044

**Authors:** Elmar W. Tobi, Catarina Almqvist, Anna Hedman, Ellika Andolf, Jan Holte, Jan I. Olofsson, Håkan Wramsby, Margaretha Wramsby, Göran Pershagen, Bastiaan T. Heijmans, Anastasia N. Iliadou

## Abstract

**Study Question:** What is the relationship with assisted reproductive technology (ART) and DNA methylation (DNAm) patterns in cord-blood and is there any difference between in vitro fertilisation (IVF) and intra-cytoplasmic sperm injection (ICSI)?

**Summary Answer:** We identify nineteen CpGs at which DNAm was associated with being conceived via assisted reproductive technology (ART). No difference was found between IVF and ICSI.

**What is known already:** Prior studies on either IVF or ICSI show conflicting outcomes as both widespread effects on DNAm and highly localized associations have been reported. No study on both IVF and ICSI and genome-wide neonatal DNAm has been performed.

**Study design, size, duration:** We measured 87 infants conceived with in vitro fertilisation (IVF) or intra-cytoplasmic sperm injection (ICSI) and 70 who were not in a cross-sectional study.

**Participants/materials, setting, methods:** Participants were from the UppstART study, which were recruited from fertility and reproductive health clinics, and the Born into Life cohort, which is created from the larger LifeGene study. We measured DNAm from DNA extracted from cord blood collected at birth with the Illumina Infinium HumanMethylation450k BeadChip micro-array. Group differences in DNA methylation at individual CpG dinucleotides (CpGs) were determined using robust linear models and post-hoc Tukey’s tests.

**Main results and the role of chance:** We found no association with global methylation levels, imprinted loci and meta-stable epialleles. In contrast, we identify nineteen CpGs at which DNA methylation was associated with being conceived via ART (effect estimates: 0.5-4.9%, P_FDR_<0.05), but no difference was found between IVF and ICSI. The associated CpGs map to genes related to brain function/development or genes connected to the plethora of conditions linked to subfertility, but functional annotation did not point to likely functional consequences.

**Limitations, reasons for caution:** We measured DNAm in cord blood and not at later ages and/or other tissues and findings remain to be replicated in an independent study.

**Wider implications of the findings:** We find ART is associated with DNA methylation differences in cord blood, but these differences are limited in number, effect size and with unknown functional consequences in adult blood. We did not find indications for a disparity between IVF and ICSI.

**Study funding/competing interest(s):** The authors declare no competing interests. The funders had no role in the study, study design, analysis and decision to publish.

## Introduction

The usage of assisted reproductive technology (ART) is increasing world-wide and its possible health consequences are a topic of intense study. Evidence is building for an association of ART with long-term health outcomes for the offspring, including autism spectrum disorders (Liu *et al.*, 2017) and cardiovascular health (higher blood pressure, suboptimal cardiac diastolic function and vessel thickness) (Guo *et al.*, 2017). A body of evidence links ART to short term health effects, including low birth weight, placenta associated anomalies (Vermey *et al.*, 2019) and pregnancy complications (Qin *et al.*, 2016), congenital malformations and imprinting disorders (Turkgeldi *et al.*, 2016). However, it is still debated if these associations stem from the application of ART or the underlying infertility leading couples to ART (Luke *et al.*, 2016). Animal studies provided potential molecular mechanisms for these observations by highlighting that the ART procedure may induce changes to epigenetic marks (Morgan *et al.*, 2008; Wang *et al.*, 2010). Epigenetic marks, such as DNA methylation (DNAm), influence the transcription potential of genomic regions (Jaenisch and Bird, 2003). Human studies likewise point towards a link between early development, DNAm and (late-life) phenotypes, but the causality of these associations remains unknown (Tobi *et al.*, 2018).

To date, several studies on DNAm and ART have been performed. Multiple studies have focused on candidate gene regions and/or global methylation levels (Lazaraviciute *et al.*, 2014; Canovas *et al.*, 2017). Genome-wide efforts have focused on samples taken from extra-embryonic lineages (Xu *et al.*, 2017; Choufani *et al.*, 2018) and when focused on material from the infant itself suffered from a small sample size (Melamed *et al.*, 2015) or batch effects correlating with ART status (Estill *et al.*, 2016). Larger studies have focused on specific ART techniques, namely intra-cytoplasmic sperm injection (ICSI) (El Hajj *et al.*, 2017) or in vitro fertilisation (IVF) (Castillo-Fernandez *et al.*, 2017) or IVF and less invasive gamete intra-fallopian transfer (GIFT) coupled with intra-uterine insemination (IUI) (Novakovic *et al.*, 2019). It has been hypothesized that the technique used may matter for the possible consequences on DNAm patterns (Loke and Craig, 2016). Overall, these studies report conflicting outcomes, with two reporting wide-spread associations between ART and (cord-)blood DNAm at birth (El Hajj *et al.*, 2017) which appear to fade with age (Novakovic *et al.*, 2019), while another study reported a DNA methylation difference at a single genomic locus only (Castillo-Fernandez *et al.*, 2017).

We undertook an epigenome wide association study on ART, comparing DNAm patterns in cord blood of children conceived via ART (N=87) (Iliadou *et al.*, 2019) with that of children from medically unassisted conceptions (henceforth “MUC”, N=70) (Almqvist *et al.*, 2011). Since we had detailed information on the ART technique used for 77 of the children conceived via ART we investigated possible different outcomes for IVF and ICSI on DNAm which has been hypothesized to be important (Loke and Craig, 2016), but is yet to be tested. In addition, we explored the possible functional consequences of these methylation differences using external datasets.

## Methods

### Study Subjects

#### ART controls: Born into Life

The prospective longitudinal birth cohort study Born into Life was originally created from the larger LifeGene study (Almqvist *et al.*, 2011). Between the years 2010 and 2012, pregnant women who were already participating in the LifeGene study and living in Stockholm County were recruited to Born into Life (Smew *et al.*, 2018). The inclusion criteria were that they had responded to baseline questionnaires from the LifeGene study, were pregnant, and gave informed consent. They were recruited both before and after 10-14 gestational weeks, but no later than 26-28 weeks. Originally 107 pregnant women were included in Born into Life. We only included the 77 women for which cord blood had been successfully collected in this study.

Women in the Born into Life were asked to complete questionnaires regarding pregnancy, lifestyle and health at 10-14 and 26-28 gestational weeks. Data on maternal self-reported smoking during pregnancy and body mass index (BMI), in kg/m2 obtained at the first antenatal care visit in gestational week week 5-12 were collected from birth records. Data regarding highest attained educational level, ranging from mandatory secondary school to high school, university or other, were retrieved from the baseline LifeGene questionnaires. Maternal age at delivery, in years, was calculated from mothers’ date of birth. Parity and data regarding the infants’ sex, gestational age in weeks and mode of delivery, defined as vaginal delivery or Caesarean section, were also collected from the birth records. Cord blood and placenta samples were obtained from delivery. Cord blood was aspirated 2 minutes after birth by the assisting midwife into a test tube and kept in room temperature awaiting transportation. The birth records regarding both mother and child at delivery were collected from Danderyd Hospital, Stockholm.

#### ART cases: UppstART

The UppstART study has been described in detail elsewhere (Iliadou *et al.*, 2019). Participants of the UppstART study were recruited from three of the four fertility and reproductive health clinics in Stockholm (one public, two private) and one private clinic in Uppsala county, which also serves a large volume of patients from Stockholm. Recruitment took place from September 2011 to December 2013. IVF treatment(s) of the participants were followed until December 2014 or drop-out/consent withdrawn (n=4), which ever came first. The participants were asked to answer a web-based baseline questionnaire within a few days of their clinic visit and prior to their IVF treatment start, which included an extensive list of questions on sociodemographic, anthropometric and life-style factors. Once the IVF treatment began and the participants reached the stage of oocyte retrieval, they were asked to respond to a second online follow-up questionnaire, with a shorter version of the baseline questionnaire, to identify any changes in lifestyle factors since the initiation of their treatment. Staff at seven delivery units in Stockholm and Uppsala were recruited to assist in collecting samples from UppStART participants during delivery. Delivery clinics were provided with a sample collection kit including tubes for collection of cord-blood for DNA extraction (EDTA). Samples were stored in −20°C freezers at the delivery units until they were collected by UppStART study staff and deposited into the KI Biobank.

### DNA methylation measurements

Genome-wide DNA methylation data were generated using the Illumina Infinium Human Methylation 450K BeadChip (450k array). A total of 500ng of genomic DNA isolated from cord blood was bisulfite treated using the EZ-96 DNA methylation kit (Zymo Research, Orange County, USA). We used the D-optimum criterion to assign samples over two 96-well plates and individual 450k arrays, ensuring even distributions of ART cases and controls, ART methods (IVF or ICSI), gender, gestational age and birth month across the two 96-well bisulfite plates and each 450k array. The 450K arrays were measured at ServiceXS (Leiden, The Netherlands). The quality of the generated 450K array data was assessed using both sample dependent and sample independent quality metrics using the *Bioconductor* package *MethylAid* (van Iterson *et al.*, 2014) with default settings. We used the *Bioconductor* package *omicsPrint* (van Iterson *et al.*, 2018) to check for sample duplications and admixtures and sample sexes were checked using the X-chromosomal CpGs. One sample was not the correct gender and was deleted. We used principal component analysis (PCA) and hierarchical clustering on the raw autosomal beta-values to search for outliers and suspect patterns in the dataset, finding none. The cell proportions of cord blood were imputed using IDOL (Koestler *et al.*, 2016) on the “FlowSorted.CordBloodCombined.450k” reference set using the estimateCellCounts2() function in the *FlowSorted.Blood.EPIC* R package(Salas *et al.*, 2018; Gervin *et al.*, 2019). Normalization of the dataset was performed by NOOB background and color correction in combination with Functional Normalization (Fortin *et al.*, 2014) using four principal components (eigenvalue > 1). All measurements with <3 beads (0.05% of probes), < 1 intensity value (0.012% of probes) and a detection p-value >0.01 (0.11% of CpGs) were set as missing. The measurement success rate per sample was >99%. CpGs with probes that did not map to unique genomic locations (Chen *et al.*, 2013) or with a < 95% measurement success rate (0.23% of CpGs) were then removed. We used custom scripts that add on the functions from the *minfi* package (Aryee *et al.*, 2014) and implements parallelization where possible (https://github.com/molepi/DNAmArray).

Outlier detection was performed by PCA and hierarchical clustering on the autosomal beta-values. Hierarchical clustering identified one possible outlier. This individual had a very low gestational age (30 weeks), the lowest in the dataset, and this clustering is therefore likely based on biological reasons. High-quality DNA methylation data were obtained for 157 individuals, 70 MUC from Born into Life, 6 ART from Born into Life (specific type of ART unknown), 81 ART from UppStART (specific type of ART unknown for N=4) for a total of 441,836 autosomal CpGs. The datasets generated and analyzed during the current study are not publicly available due to Swedish privacy and data safety laws but are available from ANI and CA on reasonable request and after meeting legal requirements.

#### Measures of global methylation

Genome-wide average DNA methylation (GWAM) (Li *et al.*, 2018) was calculated by averaging all beta-value measurements across the autosomes for each individual. The *R* package *REMP* (Zheng *et al.*, 2017) was used to infer DNA methylation at either /ALL/ or *LINES-*1 sequences and then the average across all *ALU* or *LINES-*1 elements was calculated for each neonate.

#### TFBS enrichment

We calculated transcription factor binding site enrichments using the R package *PWMEnrich* (Stojnic and Diez, 2018) using binding motifs from motifDB. We calculated position weight matrices using the DNA base background frequencies calculated for the CpGs tested with 25bp flanking sequences. Enrichment was tested relative to this background.

#### Statistics

We compared unassisted and ART cohort descriptives via Kruskal Wallis (maternal BMI, maternal SES, years till index pregnancy, CD4T, NK and nucleated red blood cells), t-test (maternal and gestational age, birthweight, CD8T, Monocytes, granulocytes and B cell proportions) assuming equal or unequal variance where appropriate and chi-square tests (child sex, parity and maternal smoking). DNA methylation was always analyzed as beta-value which is reported as % (e.g. ×100) in Tables. GWAM, *ALU* and *LINES-*1 methylation levels were compared via t.test (unassisted vs ART) or ANOVA (medically unassisted fertilization versus IVF versus ICSI).

We used the R package *cate* (Wang and Zhao, 2015) to identify latent variables (e.g. “hidden variables”) influencing the relation between DNA methylation and ART (yes/no) or ART method (IVF/ICSI/unassisted). Two latent variables were identified (P<0.001) for ART and four latent variables for ART method (P<0.001). In both cases the first two latent variables correlated with the proportion of nucleated red blood cells (nRBC) in blood (rho>0.94, P<0.001) and the other two latent variables correlated with CD4T or B cell proportions in blood and several batch effects (rho> |0.3|, P<0.001) arguing together with directed acyclic graphs (DAG) (*Supplemental text*), for a basic model adjusting for batch effects and imputed cell proportions.

We performed epigenome-wide association analyses (EWAS) on being conceived by ART or not using robust linear regression (rlm) from the R *MASS* package with White’s estimator for robust standard errors as implemented in the R package *sandwich* (Zeileis, 2006) (which leads to a model robust for outlying beta-values and heteroscedasticity). To test for differences in neonates conceived via IVF or ICSI or without medical assistance we used type II ANOVA with White’s estimator for robust standard errors to account for any heteroscedasticity and unequal variances between groups using the R *car* package. We employed heteroscedasticity robust Tukey tests to test for DNA methylation differences between IVF, ICSI and unassisted conceptions post-hoc. In all instances we used the beta-value of an individual CpG dinucleotide as the outcome and ART status as the dependent. We adjusted for the sex of the individual, height of the sample on the 450k array glass slide (continues variable from 1-6), scan batch and bisulfite plate and the imputed cell proportions (CD4T, CD8T, Mono, NK, Bcell, Gran, nRBC). We ran additional sensitivity analyses by omitting the cell proportions or expanding the adjustment with minimal adjustment sets coming from DAG analysis with the following models, i: years till the index pregnancy, maternal age, pre-pregnancy BMI, educational attainment as proxy for socioeconomic status, smoking history ii) gestational length and parity. In addition, we performed look-ups in other published meta-EWAS of prenatal exposures.

Regions were tested with the same adjustments as defined above in a linear mixed model with a random effect for individual and a factorial co-variate denoting the specific CpG dinucleotide of each DNAm measurement in the R *lme4* package using compound symmetry as correlation structure and the R *ImerTest* package for the Satterthwaite’s approximation of the degree of freedom of the fixed effects within each linear mixed model.

All analyses were performed in R (version 3.6.1) (R core team, 2019). All p-values reported are two-sided and multiple testing correction is done using FDR (false discovery rate).

#### Ethical approval

Ethical approval for Born into Life was granted by the Regional Ethics Review Board in Stockholm, Sweden. Informed consent from both parents was obtained for all study participants. The UppstART study has been approved by the regional ethics review board at the Karolinska Institutet (Dnr 2011/23031/1, Dnr 2011/1427-32, Dnr 2012/131-32, Dnr 2012/792-32, 2013/1700-32, 2014/1956-32, Dnr 2015/1604-32). Women and their partners were approached by the clinic nurse and asked to participate in the study. To facilitate the process of informed consent, the couple was provided with information approved by the regional ethical board, both verbally and in written format about the purpose of the study, methods, possible risks, and that participation was voluntary. Additionally, participants were informed that they could withdraw from the study at any time with no impact to their medical care. The requirement for inclusion in the study was an understanding of the Swedish language and exclusion was the use of donor gametes.

## Results

### Study subjects

Pre-processing and normalization resulted in DNAm data of 157 infants from cord blood, 87 of which were conceived with the help of ART (Table 1). We had detailed information on the ART technique used for 77 of the 87 newborns conceived by ART and with DNAm data, of which 44 were conceived via IVF and 33 via ICSI. The mothers that conceived with the help of ART were not different in body mass index (BMI, P=0.25) and age (P=0.25), but had a lower socio-economic status (P<0.001) and more were former smokers (P<0.001) than mothers with a MUC. It took on average two years longer for the mothers who conceived through ART to become pregnant than for mothers with MUC (P<0.001). There was no difference between ART and MUC in length of gestation (P=0.16) and parity (P=0.20). The percentage of male newborns in the MUC group was higher (61.4% vs 50.6%), although this difference was not significant (P=0.23).

**Table 1.** Cohort characteristics

| Variable | MUC<sup>#</sup> | ART | IVF | ICSI |
| --- | --- | --- | --- | --- |
| N | 70 | 87 | 44 | 33 |
| Frozen (N)<sup>1</sup> | - | 10 | 7 | 3 |
| Blastocyst (N)<sup>2</sup> | - | 8 | 5 (3 frozen) | 3 (2 frozen) |
| BMI in kg/m<sup>2</sup> (SD) | 22.8 (3.5) | 23.4 (3.4) | 23.5 (3.3) | 23.4 (3.0) |
| Age in y(SD) | 32.4 (3.6) | 33.1 (3.6) | 33.0 (3.7) | 33.0 (3.7) |
| SES<sup>3</sup> (SD) | 3.0 (0.3) | 2.6 (0.8)<sup>*</sup> | 2.7 (0.7) | 2.3 (0.9) |
| Smoking |  |  |  |  |
| Never | 65 | 56 | 27 | 23 |
| Ever | 4 | 26 | 17 | 9 |
| NA | 1 | 5 | 0 | 1 |
| Years to index pregnancy |  |  |  |  |
| Mean (SD) | 0.2 (0.7) | 2.6 (1.3)<sup>*</sup> | 2.7 (1.2) | 2.4 (1.5) |
| Min - max | 0- 4 | 0 – 7 | 1- 7 | 0-7 |
| Parity |  |  |  |  |
| First | 47 | 62 | 34 | 22 |
| Second | 21 | 17 | 10 | 7 |
| Third | 2 | 1 | 0 | 1 |
| NA | 0 | 7 | 0 | 3 |
| N Male children (%) | 43 (61.4) | 44 (50.6) | 22 (50.0) | 16 (48.5) |
| Gestational age in weeks | 39.1 (1.6) | 39.5 (2.0) | 39.2 (2.3) | 39.7 (1.6) |

1. The number of embryos that were frozen and thawed before placement *in utero*. 2. The number of embryos that were grown *ex utero* to the blastocyst stage before placement *in utero*. Off note, five of these blastocysts were frozen and thawed. 3. Socio-economic status (SES) graded on a 4 level scale based on the highest attained educational level. - <sup>\*</sup>Significantly different P<0.001, <sup>#</sup> medically unassisted conception

**Table 2.**
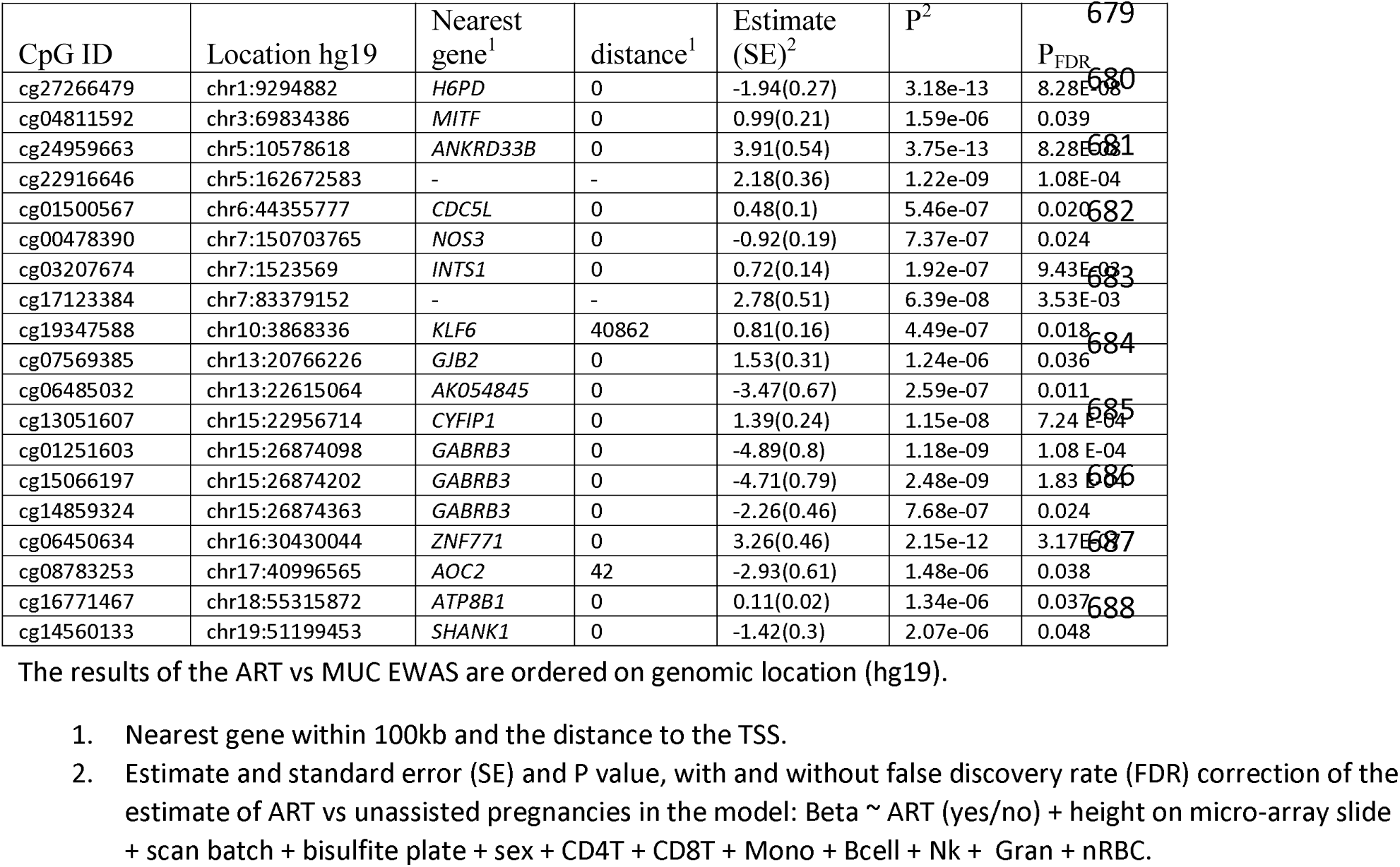
Results of ART EWAS

DNAm is a key determinant of cell identity (Jaenisch and Bird, 2003) and cord blood consists of multiple cell types that influence DNAm (Jones *et al*, 2007). Seven major cell types, including nucleated red blood cells, were imputed from the genome-wide DNAm data of the newborns (Gervin *et al*, 2019). In concordance with a prior report on unassisted and ICSI newborns (El Hajj *et al.*, 2017), there were no differences in these seven cell proportions between ART and MUC cord blood samples (P_nominal_>0.13, Supplemental Figure S1).

### Global DNAm comparison between ART and unassisted conceptions

First, we investigated genome-wide average DNA methylation (GWAM) (Li *et al*, 2018) in the cord blood of neonates conceived via IVF or ICSI or MUC. There was no difference between these 3 groups (P=0.89, Supplemental Figure S2). Next, we investigated the genome-wide average DNA methylation of ALU and LINES-1 elements, an often used proxy for global DNA methylation (Zheng *et al.*, 2017), finding no difference between groups (P>0.2, Figure S3).

#### DNAm comparison between ART and medically unassisted conceptions

We compared DNA methylation at 441,836 autosomal CpGs between 87 ART and 70 MUC neonates (Supplemental Figure S4). Nineteen CpGs were associated with being conceived via ART in a model adjusting for sex, batches and cell heterogeneity (P_FDR_<0.05, Figure S5). The associations extended to neighboring probes (Figure 1) for the proximal promoter of *AK054845*, a gene of unknown function (five CpGs P_nominal_<0.05), and the proximal promoter of a smaller variant of *GABRB3* (NM_001191321.2) which is expressed in the brain (eight CpGs P_nominal_<0.05) and has a neuro-developmental role (Tanaka *et al.*, 2012). Indeed, several CpGs can be linked to genes with roles in brain function or (early) brain development. *ZNF771* (cg06450634), as a transcription factor (TF), is a driving factor behind a large gene network in the brain (Maulik *et al.*, 2018) while *CYFIP1* (cg13051607) (Hsiao *et al.*, 2016), *GABRB3* (cg01251603, cg15066197,cg14859324) (Tanaka *et al.*, 2012) and *SHANK1* (cg14560133) (Sungur *et al.*, 2018) are all autism spectrum disorder candidate genes, a late life phenotype tentatively associated with ART (Liu *et al.*, 2017).

**Figure 1.**
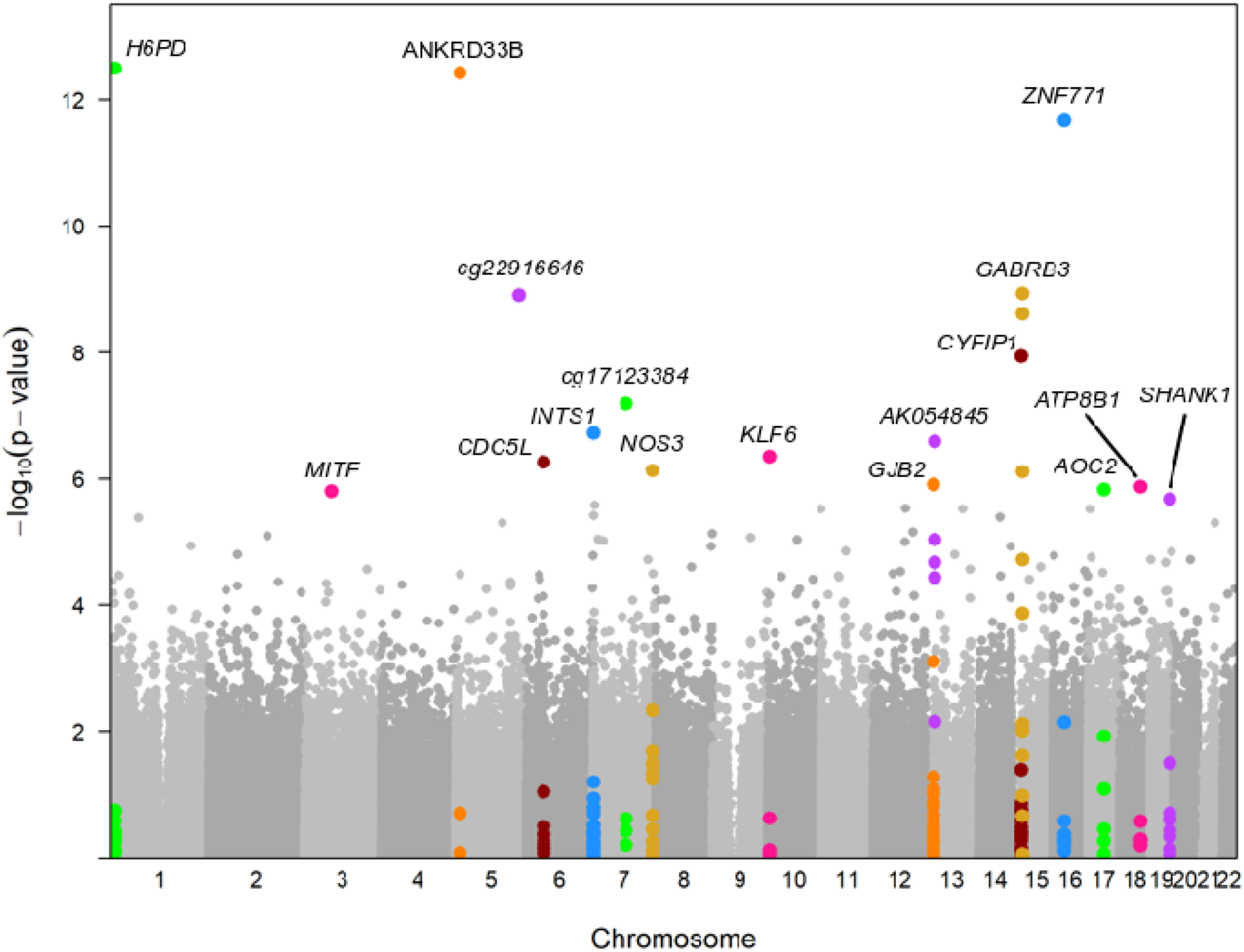
A Manhattan plot showing the –log10 P-values (y-axis) for the association of DNA methylation at individual CpG dinucleotides across the 22 autosomal chromosomes (x-axis). CpG dinucleotides 5kb up- and downstream of the lead association have been colored in the same color as the lead association.

Omitting the adjustment for cell heterogeneity had little to no effect on the effect estimates for all nineteen CpGs (Supplemental Table S1) just like additional adjustment for maternal and pregnancy characteristics (Supplemental Table S2). The nineteen CpGs were not among those previously reported in other EWASs (FDR corrected P-value of <0.05) with prenatal smoking (Joubert *et al.*, 2016b), folate use (Joubert *et al.*, 2016a), maternal hypertension and pre-eclampsia (Kazmi *et al.*, 2019), maternal BMI (Sharp *et al.*, 2017) and birthweight (Küpers *et al.*, 2019). Moreover, a look-up of these previously reported CpGs in our study did not yield more nominally associated CpGs with ART than may be expected by chance (P>0.07).

#### DNA methylation comparison between IVF, ICSI and unassisted conceptions

To investigate any differences between IVF and ICSI we performed post-hoc Tukey’s tests for DNA methylation differences between neonates conceived via IVF and ICSI for these nineteen CpGs. No differences were found (P_FDR-19tests_>0.15). Next, we extended our analysis to all CpGs by performing an ANOVA analysis to test for differences in DNA methylation across the 441,836 autosomal CpGs between neonates conceived with IVF (N=44) or ICSI (N=33) or MUC (N=70). This yielded five CpGs already identified in the EWAS for ART status and in all cases the difference was between the MUC and both IVF and ICSI, thus finding no evidence for IVF or ICSI specific associations.

#### Functional annotation

We performed KEGG and Gene Ontology tests (Phipson *et al.*, 2015), finding no enrichment for the genes linked to nineteen CpGs associated with ART, nor when we relaxed the significance threshold to suggestive associations (P<10^−5^, N=37). We did not find significant enrichments of single transcription factor (TF) binding motifs (Stojnic and Diez, 2018), although multiple CpGs overlapped TF binding sites of TFs with a role in early development like *TFAP2C* (Sharma *et al.*, 2016) overlapping cg27266479 mapping to *H6PD* and *RAX* (Bennett *et al.*, 2008) overlapping cg16771467 to *ATP8B1*. Look-up of the nineteen CpGs in reference data (Bonder *et al.*, 2017) uncovered no link between DNAm and gene-expression in adult blood. We measured DNAm in cord blood and average DNAm levels may vary between tissues, but still reflect variation in other tissues (Slieker *et al.*, 2013). There was little to no correlation of methylation at the nineteen CpGs between adult whole blood and tissues from sixteen cadavers (Slieker *et al.*, 2013; Relton *et al.*, 2015), with the notable exception of cg14560133 (*SHANK1*, β = -1.42% (SE=0.3%), P=2.1×10^−6^) which showed moderate correlation with 5/8 tissues (rho>0.53, P<0.04), Figure 2). The three CpG dinucleotides at *GABRB3* (cg01251603, cg15066197,cg14859324), a gene active in the brain, showed weak to moderate correlation between adult blood and four brain regions (rho=0.39-0.62, P<0.001, N=71-74) in another reference dataset (Hannon *et al.*, 2015).

**Figure 2.**
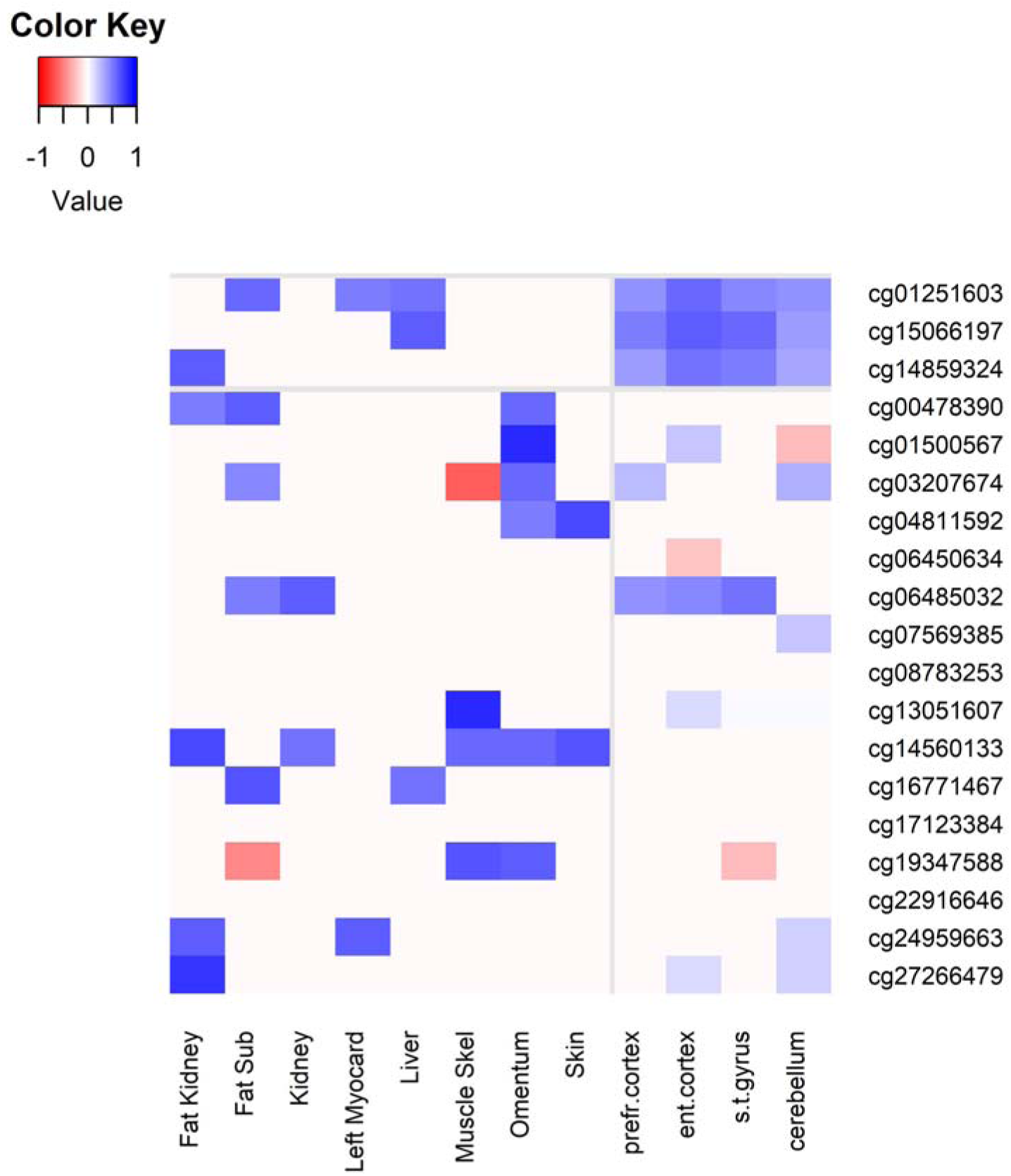
The spearman correlation plotted for 8 tissues from the body (N=16, P<0.1) and 4 from the brain (N>71, P<0.01) in colors ranging from bright red (*rho*= -1.0) to dark blue (*rho*=1.0). The light gray lines along the rows demark three CpG dinucleotides from the *GABRB3* region associated with ART. The light gray line running along one column denotes the separation of the two tissue reference datasets used. *“Fat sub”: subcutaneous fat, “Left myocard”: left myocardium, “Muscle Skel”: skeleton muscle, “prefr. Cortex”: prefrontal cortex, “ent.cortex”: entorhinal cortex “s.t.gyrus”: superior temporal gyrus*.

#### Imprinted regions and metastable epialleles

Multiple candidate gene studies have looked at DNA methylation or gene-expression differences of imprinted genes between ART and MUC with mixed results. Therefore, we looked at 374 CpGs on the 450k array known to overlap imprinted differentially methylated regions (DMRs) (Yuen *et al.*, 2011). There was no overlap with the CpGs showing a (suggestive) association with ART in our study (P<10^−5^, N=22). These 374 CpGs where distributed over 59 regions. There was no difference in DNAm between MUC and ART infants when we tested each of these 59 regions (P_FDR_>0.86). Similarly, so-called metastable epialleles (MEs) are hypothesized to be especially sensitive to the early prenatal environment (Kessler *et al.*, 2018). There were 187 CpGs overlapping MEs, none of which were associated with ART (P_FDR_>0.06).

#### Prior studies

We performed a look-up of CpGs and regions associated in prior studies with one form of ART. The sole region associated with IVF in the meDIP sequencing study from Castillo-Fernandez et al. was not covered by our genome-scale screen with the 450k array (Castillo-Fernandez *et al.*, 2017). Next, El Haij et al. found 4730 CpG dinucleotides associated with ICSI after FDR adjustment (which are unavailable as Supplemental information) of which two regions, out of five regions selected for validation with pyrosequencing, were validated in an additional sample (El Hajj *et al.*, 2017). The seven CpGs in these two regions which cover the proximal promoters of *ATG4C* and *SNORD114-9* were not associated with ART in our study (P>0.23) nor with ICSI only (post hoc Tukey’s test: P>0.40).

Using the 850k array, Novakovic et al. identified 2340 CpG dinucleotides at which DNA methylation in Guthrie card blood was associated with IVF and GIFT coupled with IUI (Novakovic *et al.*, 2019), 1228 of which were covered in our 450k study. Of these, 38 were nominally associated (P<0.05) and with the same direction of effect in our study. This decreased to 12 loci when we tested for DNA methylation differences between MUC and IVF only. Novakovic et al. also identified three differentially methylated regions where the difference in methylation associated with IVF remained in an adult sample. CpG dinucleotides at *CHRNE* (chr17:4803506-4805392), *PRSS16* (chr6:27185896-27186199) and *TMEM18* (chr2:731073-732037) showed a similar direction of effect in our study as that of Novakovic et al. and the association was almost nominally significant for *PRSS16* (cg10279314: β=2.3%[SE=1.2%], P=0.057, cg09395805: β =3.1[SE=1.6],P=0.049, cg07555084: β =2.2[SE =1.2],P=0.074).

## Discussion

We identified nineteen CpGs at which DNAm was associated with ART and the association extended to neighboring CpG dinucleotides at *AK054845* and *GABRB3*. We found no indication for a disparity between IVF or ICSI on DNAm patterns. The associations were robust to adjustment for cellular heterogeneity and maternal characteristics and did not overlap with those loci associated with other common prenatal exposures. We found no evidence that imprinted loci and meta-stable epialleles are especially sensitive to ART. The nineteen CpGs can be annotated to relevant genes, but the functional implications of variation in DNA methylation at these CpGs remains to be elucidated.

The nineteen CpG dinucleotides could be mapped to fifteen genes. Four of the nineteen were located in a proximal promoter and here the neighboring CpG dinucleotides were also nominally associated with ART. In other cases the associations were either limited to one CpG dinucleotide, which may stem from the sparse coverage of the 450K array across the methylome and in other cases is consistent with the fact that DNAm acts through the altering of the binding potential of a specific TFBS (Bonder *et al.*, 2017). Indeed, none of the CpGs were located in a CpG island, which is consistent with data from other exposures showing that associations between DNAm and prenatal and/or environmental conditions are enriched at regulatory regions like enhancers and CpG poor promoters that show intermediate levels of methylation (Tobi *et al.*, 2014).

Several of the CpGs can be linked to genes with a role in brain function and/or development. *ZNF771* (cg06450634) has been identified as a driving factor behind a large gene network in the brain (Maulik *et al.*, 2018) and *CYFIP1* (cg13051607) (Hsiao *et al.*, 2016), *GABRB3* (cg01251603, cg15066197,cg14859324) (Tanaka *et al.*, 2012) and *SHANK1* (cg14560133) (Sungur *et al.*, 2018) are all autism spectrum disorder candidate genes, a late life phenotype tentatively associated with ART (Liu *et al.*, 2017). In addition, several CpG dinucleotides can be linked to genes connected to the plethora of conditions that can be linked to subfertility. *H6PD* (cg27266479) is a candidate gene for polycystic ovary syndrome (PCOS) (Martinez-Garcia *et al.*, 2012). *INTS1* (cg03207674) has a crucial role in the developing blastocyst, as inhibition of *INTS1* function causes growth arrest (Hata and Nakayama, 2007). *NOS3* (cg00478390) knockout mice are used as an *in vivo* model of (recurrent) embryo loss, as nitric oxide metabolism plays an important role in implantation (Pallares and Gonzalez-Bulnes, 2010).

The latter may hint that the various medical reasons for ART may underlie the associations, rather than the ART process itself, which is a key question in the study of ART (Luke *et al.*, 2016). We employed directed acyclic graphs (DAG) (Krieger and Davey Smith, 2016) and DAG indicated that and ascertainment of a direct effect is possible with a small minimal adjustment set. There was little to no effect on the effect estimates from adjustment for various maternal characteristics, gestational age and the number of years until pregnancy (as a proxy for in-/subfertility). In addition, we did not find any evidence for different or stronger effects of ICSI on DNAm, as has been hypothesized (Loke and Craig, 2016), and the fertility clinics from which we recruited all used the same culturing media for ICSI and IVF. The last two items highlighted might argue that not the (reasons for) infertility, but the ART process itself may explain the associations.

Our analysis did not find wide-spread genome-wide differences like other genome-scale studies to date (El Hajj *et al.*, 2017; Novakovic *et al.*, 2019), but is consistent with a genome-scale study on only IVF using immunoprecipitation of methylated DNA with next generation sequencing (Castillo-Fernandez *et al.*, 2017). We extensively compared individual results, as far as possible, and found little overlap. This may be due to study power, the different scope of the measurement techniques used (Castillo-Fernandez *et al.*, 2017) and the different sources of genomic DNA (Guthrie card versus whole cord blood) (Novakovic *et al.*, 2019). In addition, it is possible that different media was used between countries or that the fact that only a small proportion of our IVF and ICSI group consisted of embryos that were frozen and/or cultured extensively in comparison to other studies. The influence of this latter aspect we were underpowered to test as most ART procedures entailed fresh embryos that were not cultured extensively *in vitro*.

Our study is one of the larger genome-scale DNAm studies on ART to date and we are the first to study both IVF and ICSI on this scale, nonetheless our study has important limitations to consider. First, DNAm is one of the drivers of cell identity (Jaenisch and Bird, 2003) and we measured cord blood, which may be a tissue less relevant in relation to study outcomes although it may still mark processes in relevant tissues due to mitotic inheritance (Heijmans and Mill, 2012). We used the latest methods to impute the seven major cell types in cord-blood (Gervin *et al.*, 2019) and in conjuncture with an earlier report (El Hajj *et al.*, 2017) these imputed cell types did not differ between infants conceived with or without ART and adjustment did not alter the effect estimates. However, most of these major cell types consist of subsets of more specialized cells (subsequently showing unique DNAm profiles at increasingly select loci) and their influence could not be investigated. However, none of the nineteen CpG dinucleotides is linked to a gene with a role in (auto)immune function or hematopoiesis. Another major driver of DNAm variation is genetic variation (Bonder *et al.*, 2017) and we did not control for genetic variation in our analyses. Nine out of nineteen CpG dinucleotides did not have a single nucleotide polymorphism (SNP) influencing the DNA methylation levels, as determined in a large meta-analysis (Bonder *et al.*, 2017) and none of these 450k probes had an overlap with a single SNP arguing that genetic heterogeneity is unlikely to underlie our findings. In addition, the influence of the prenatal environment on DNAm variation may be independent and additive of genetic variation (Tobi *et al.*, 2012).

Also important is to consider the fact that an ART population is inherently different from the general population (Luke *et al.*, 2016). Despite controlling for possible confounding factors, and crosschecking our results with other EWAS on prenatal complications/exposures, unresolved confounding remains a possibility. Our study differs with some of the earlier described in ART studies, which is useful for triangulation between studies (Lawloret *al*., 2017), as our control group has a higher, rather than a lower, SES as is normal in most of studies on ART.

The data presented showcases modest and specific DNAm differences associated with ART, of which the functional relevance in adult tissues is unknown. We did not find any difference in DNAm patterns between IVF and ICSI. Our study found little evidence for the hypothesis that ART, be it IVF or ICSI, leads to wide-spread disturbances of DNAm patterns nor for the hypothesis that ICSI has a different/larger relation with DNAm patterns. Our findings warrant cautious interpretation and awaits confirmation, given the sample size, tissue studied and the unknown functional consequences of the identified DNAm differences.

## Data Availability

Access to the Data is available upon request

## Author contributions

Conceptualization: ANI, CA. Methodology: EWT, BTH. Investigation: EWT, BTH, ANI, CA. Formal Analysis: EWT. Validation: EWT. Resources: ANI, AH, CA, GP, EA, JIO, MW, HW. Data curation: ANI, AH. Writing - original draft: EWT. Writing - Review & Editing: BTH, ANI, AH, CA, GP, EA, JIO, MW, HW. Visualization: EWT. Supervision: BTH, ANI. Project administration: ANI, AH. Funding Acquisition: ANI, CA, GP, EA

## Acknowledgements

We wish to thank the Biobank at the Karolinska Institutet for professional biobank service.

## Competing interests

The authors declare no conflicts of interest

**Abbreviations**
ART: assisted reproductive technology
IVF: *in vitro* fertilisation
ICSI: intra-cytoplasmic sperm injection
DNAm: DNA methylation
EWAS: epigenome-wide association analysis
CpGs: CpG dinucleotides
IUI: intra-uterine insemination
GIFT: gamete intra-fallopian transfer
MUC: medically unassisted conceptions
450K array: Illumina Infinium Human Methylation 450K BeadChip
PCA: principal component analysis
TFBS: transcription factor binding site
TF: transcription factor
SNP: single nucleotide polymorphism
SES: socio-economic status
BMI: body mass index

